# Protection after Quarantine: Insights from a Q-SEIR Model with Nonlinear Incidence Rates Applied to COVID-19

**DOI:** 10.1101/2020.06.06.20124388

**Authors:** Jose Marie Antonio Minoza, Jesus Emmanuel Sevilleja, Romulo de Castro, Salvador E. Caoili, Vena Pearl Bongolan

## Abstract

Community quarantine has been resorted to by various governments to address the current COVID-19 pandemic; however, this is not the only non-therapeutic method of effectively controlling the spread of the infection. We study an SEIR model with nonlinear incidence rates, and introduce two parameters, *α* and *ε*, which mimics the effect of quarantine (*Q*). We compare this with the Q-SEIR model, recently developed, and demonstrate the control of COVID-19 without the stringent conditions of community quarantine. We analyzed the sensitivity and elasticity indices of the parameters with respect to the reproduction number. Results suggest that a control strategy that involves maximizing *α* and *ε* is likely to be successful, although quarantine is still more effective in limiting the spread of the virus. Release from quarantine depends on continuance and strict adherence to recommended social and health promoting behaviors. Furthermore, maximizing *α* and *ε* is equivalent to a *50%* successful quarantine in disease-free equilibrium (DFE). This model reduced the infectious in Quezon City by *3.45%* and Iloilo Province by *3.88%*; however, earlier peaking by nine and 17 days, respectively, when compared with the results of Q-SEIR.

## INTRODUCTION

Preventive and control measures such as hand hygiene, community quarantine and social distancing have been widely used to limit the spread of COVID-19 (Güner et al., 2020). In the Philippines, an enhanced community quarantine (ECQ) was implemented on March 16 (Official Gazette, 2020). Experts and policy makers continuously discuss what areas should or need to remain under various versions of quarantine. Quarantine is used to lower the number of infectives, helping health facilities cope, but trade-offs with the economy can be observed. A report by Jackson et al. (2020) stated that the COVID-19 pandemic is having a noticeable impact on global economic growth, such that estimates so far indicate the virus could trim global economic growth by as much as *2.0%* per month if current conditions persist. Global trade could also fall by *13%* to *32%*, depending on the depth and extent of the global economic downturn. Findings by Ozili and Arun (2020) show that the increasing number of lockdown days, monetary policy decisions and international travel restrictions severely affected the level of economic activity and stock prices. The maximum impact will not be known until the end of the pandemic.

Previously, Bongolan et al. (2020) modified the standard SEIR model with a quarantine, Q(t), parameter. In this paper, we explore post-quarantine scenarios. We consider the SEIR model with Crowley–Martin incidence rate to describe the effects of contact rate *β*, social awareness rate α among susceptibles and magnitude of interference ε among infectious populations. In this study, we generalize the notion of α and ε to represent combinations of behavioral factors and disease-resistance factors that disrupt transmission, with α and ε applying to S and I, respectively.

## METHODOLOGY

### Model Formulation

Recent studies (Zhou and Cui, 2011; Upadhyay et al., 2019) have developed models for epidemiology and virus dynamics with the Crowley-Martin incidence rate 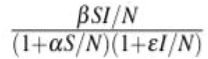. Here, we combined the Q-SEIR epidemiology model with the Crowley-Martin incidence rate (non-linear). The following were the assumptions made:

1. The entire population *N* is divided into four compartments: susceptible, *S* (those at risk of being infected), exposed, *E* (latently infected but not yet infectious), infectious, *I* (capable of transmitting the virus), and removed, *R* (removed either by recovery with permanent immunity or death). Thus, we have *N = S +E + I + R*.
2. All compartments of SEIR are well-mixed and interact homogeneously with each other.
3. Disease is transmitted from the infectious to the susceptible population by the Crowley-Martin incidence rate 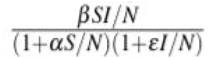. Here, *β* is the contact rate due to the infectious population, while *α* and *ε* are the inhibition effects due to susceptible and infectious populations, respectively.
4. *σ* and *γ* are invariant properties of the host-virus pair while α and ε are properties of the populations *S* and *I*, respectively.
5. From the previous model, Bongolan et al. (2020), quarantine is lifted starting May 16. (Q(t) = 1).

From the schematic diagram presented in Figure 1, the transmission process is formulated by the following system of differential equations:

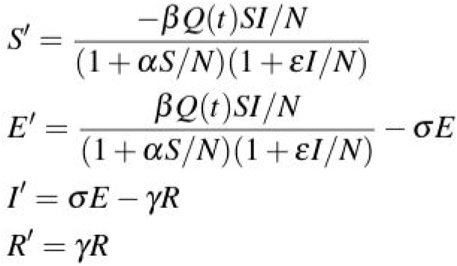

with initial conditions S(0) > 0, E(0) > 0, I(0) > 0, and R(0) > 0. All the parameters in the system are positive quantities.

**Fig. 1:**
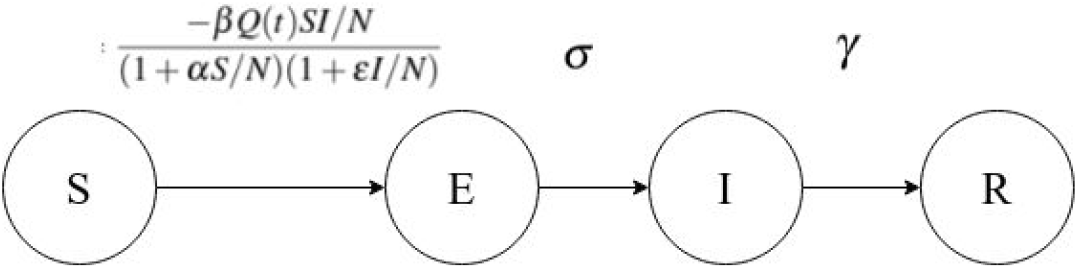
Schematic Diagram for Q-SEIR model with Nonlinear Incidence Rate

### Reproduction Number and Sensitivity Analysis

The epidemiological definition of *R*_*0*_ is the average number of secondary cases produced by one infected individual introduced into a population of susceptible individuals, where an infectious individual has acquired the disease, and susceptible individuals are healthy but can acquire the disease. (van den Driessche, 2017; Delamater et al., 2019; Hartfield and Alizon, 2013).

Using next-generation matrix method (Diekmann et al, 2010; van den Driessche, 2017), the derived reproduction number is

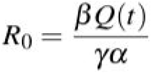

This is considered the generalized *R*_*0*_ for the model and with the assumption of post lockdown scenario where *Q(t) = 1, R*_*0*_ is

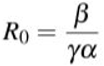

Consider post-quarantine scenario and *ε1 = 1 - ε* as the fraction of *I* considered as asymptomatic and might be under isolation, they could be attributed for mild spread of the virus. It can also be thought of as an initial infectious class before the pathogen has fully developed in the host. If *ε1* > *ε*, members of *I* can be viewed as having low natural immunity and there is a high number of vulnerable with pre-existing medical conditions among them. From that, reproduction number can be extended to

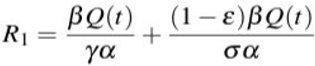

Knowledge of the relative importance of the various factors responsible for transmission is useful in assessing the best control steps. To understand the effect of the parameters, particularly *α* and *ε*, the following sensitivity analysis is conducted for the model: (1) sensitivity index of R_0_ and R_1_, (2) elasticity index (normalized sensitivity index), (3) and global sensitivity analysis (using Morris Method). The sensitivity index determines which parameters have a high impact on R_0_ and R_1_. The sensitivity index of R_i_ with respect to a parameter is 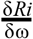 while the elasticity index (normalized sensitivity index) measures the relative change of R_i_ with respect to parameter ω, denoted by *γ*_ω_^Ri^ and defined as

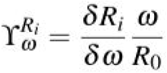

An elasticity index guides the setting of values for each parameter to control infection.

On the other hand, the Morris Method was used for global sensitivity analysis. This allows us to classify the input parameters into three groups: parameters having negligible effects, parameters having large linear effects without interactions and parameters having large nonlinear effects with or without interactions. (Iooss and Lemaître, 2015).

## RESULTS

In this section, results for the sensitivity analyses of parameters are discussed. Furthermore, we compared the estimates of the previous model (Q-SEIR) with the estimates of the SEIR with a nonlinear incidence rate for Quezon City and Iloilo Province.

### Sensitivity Analysis of the Model

#### Reproduction Number and its Sensitivity and Elasticity Indices

The reproduction numbers *R*_*0*_ and *R*_*1*_ were analyzed using the six parameters in the model. Sensitivity and elasticity indices will provide insights on the effect of the parameters and on possible control strategy for minimizing the spread of the virus.

Based on Table 1, the social awareness rate, α, is the most influential parameter in the model (i.e. *R*_*0*_ and *R*_*1*_ are most sensitive to *α*, having largest absolute values). It shows that increasing (or decreasing) *α* by 10%, *R*_*0*_ and *R*_*1*_ decrease (or increase) by 2133.33% and 7893.33%, respectively. The second most influential parameter is *Q*. Increasing (or decreasing) it by 10%, *R*_*0*_ and *R*_*1*_ increases (or decreases) by 1066.67% and 3946.67%, respectively. Interestingly, *σ* and *ε* show more sensitivity to *R*_*1*_ than to *R*_*0*_, such that increasing (decreasing) them by 10%, *R*_*1*_ will decrease (increase) by 5760% and 64% respectively.

**Table 1:** Baseline Parameter and Senstivity Indices

| Parameter | Value | Sensitivity on $R_0$ | Sensitivity on $R_1$ |
| --- | --- | --- | --- |
| $Q$ | 0.2 | 106.667 | 394.667 |
| $\beta$ | 3.2 | 6.667 | 24.667 |
| $\sigma$ | 0.1 | 0.000 | -576.000 |
| $\gamma$ | 0.3 | -71.111 | -71.111 |
| $\alpha$ | 0.1 | -213.333 | -789.333 |
| $\varepsilon$ | 0.1 | 0.000 | -6.4 |

As shown in Table 2 for the baseline parameter values, the elasticity indices suggest the possibility of decreasing the values of *Q* and *β* and increasing those of parameters *γ, α, σ*, and *ε* to decrease the basic reproduction number. As to reproduction number *R*_*1*_, it is noted that *σ* has higher relative effect with 0.7297 compared to *ε* with 0.0081. Thus, a control strategy that involves social distancing (maximizing *α*), hygienic practices (maximizing *α*), contact tracing/isolation (minimizing *σ*), improvement of health resources for faster recovery rate (minimizing *γ*), vaccination (maximizing *ε*) results in better control; and quarantine (minimizing *Q*) is very effective in limiting the spread of the virus.

**Table 2:** Baseline Parameter and Elasticity Indices

| Parameter | Value | Sensitivity on $R_0$ | Sensitivity on $R_1$ |
| --- | --- | --- | --- |
| $Q$ | 0.2 | 1 | 1 |
| $\beta$ | 3.2 | 1 | 1 |
| $\sigma$ | 0.1 | 0 | -0.7297 |
| $\gamma$ | 0.3 | -1 | -1 |
| $\alpha$ | 0.1 | -1 | -1 |
| $\varepsilon$ | 0.1 | 0 | -0.0081 |

#### Morris Method for Global Sensitivity Analysis

In the Morris method, it is assumed that the initial conditions are *S = 99.9998985, E = 0.0001, I = 0.000001, R = 0.0000005* (based on Quezon City March 2020 data). The ranges of the input parameters we used are:

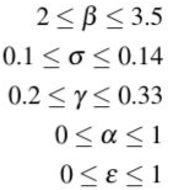

Based on Figures 2, 3, 4, *α* contributes more than ε in the dispersion of the values for *S, E* and *I*; but interestingly, *α* and *ε* show significant influence on R (shown in Figure 5). Moreover, *α* shows nonlinear and interaction effects with other input parameters. On the other hand, small perturbations on *ε* suggest a linear relationship with the values of *S, E, I* and *R*, which could be explained by the small initial value of *I*. Overall, *β, σ, γ* significantly contribute to the dispersion of the values for *S, E, I*, and *R*.

### Estimates for Quezon City

In this experiment, Quezon City (QC) is estimated to have a population of 3,240,000 in 2020. The start of quarantine was March 16; the parameters we used are: *β = 3.2, σ = 0.142857, and γ = 0.33*. For the accuracy of the estimates, we incorporated an eight day testing delay (observed range 7-10 days), thus the ground truth data on May 13 is backdated to May 5. From this, the following were derived: *Q = 0.2, α = 0.36, ε = 0.10*. Table 3 shows that, in the ideal situation (*α = 1 and ε = 1*), lifting the quarantine on May 16 predicts that *16.54%* of the population becomes infectious, down from the *19.99%* given by the original Q-SEIR model, where *α = 0* and *ε = 0*.

**Table 3:** Estimates for QC after lifting QUarantine on May 16

| $\alpha$ | $\varepsilon$ | Peaked Date | Pop. (%) |
| --- | --- | --- | --- |
| 1 | 1 | June 14, 2020 | 16.54 % |
| 0.75 | 0.25 | June 11, 2020 | 17.76 % |
| 0.5 | 0.5 | June 23, 2020 | 18.31 % |
| 0.25 | 0.75 | June 23, 2020 | 18.56 % |
| 0.25 | 0.25 | June 23, 2020 | 19.10 % |
| 0 | 0 | June 23, 2020 | 19.99 % |

### Estimates for Iloilo Province

For the estimates of Iloilo Province, it is extrapolated that the total population for 2020 is 2,524,213. This simulation has the following conditions: quarantine started March 20, *β = 2.8, σ = 0.1428571429, and γ = 0.33*. The parameter estimates were data fitted with the May 13 ground truth where 48 cases had been reported. This yielded the following values: *Q = 0.35, α= 0.32, and ε = 0.33*. Lifting quarantine on May 16, the model projects *15.34%* I compared to Q-SEIR’s *19.22%* (Table 4).

**Table 4:** Estimates for Iloilo Province after lifting QUarantine on May 16

| $\alpha$ | $\varepsilon$ | Peaked Date | Pop. (%) |
| --- | --- | --- | --- |
| 1 | 1 | July 1, 2020 | 15.34 % |
| 0.75 | 0.25 | June 26, 2020 | 16.32 % |
| 0.5 | 0.5 | June 22, 2020 | 17.33 % |
| 0.25 | 0.75 | June 18, 2020 | 17.94 % |
| 0.25 | 0.25 | June 18, 2020 | 18.28 % |
| 0 | 0 | June 14, 2020 | 19.22 % |

## DISCUSSION

Various groups (Alvarez et al., 2020; Fang et al.. 2020; Kissler et al., 2020) have demonstrated that quarantine delays COVID-19 transmission. In Bongolan et al. (2020), Q(t) was factored in to capture the effect of quarantine on the spread of the disease using the SEIR model. However, quarantine is unsustainable as it disrupts economic activity. Thus, we are modelling post-quarantine scenarios by introducing α and ε parameters. Values for α and ε were explored to understand their effects on the SEIR model as quarantine is lifted, *Q(t) = 1*. Parameters α and ε, which correspond to *β* and *γ* in Upadhyay et al. (2019), are the social awareness rate among susceptibles and the magnitude of interference within individuals in the infectious population, respectively. In practice, α and ε each represent a combination of behavioral factors (e.g., social distancing, wearing of masks, and handwashing) and disease-resistance factors (e.g., genetics, nutrition, exercise and overall health status) that disrupt disease transmission, with *α* and *ε* applying to S and I, respectively. This generalizes the interpretation of Upadhyay et al. (2019) who associated *α* only with behavior and *ε* with both behavior and disease resistance.

Using sensitivity and elasticity indices, α was found to be the most influential parameter on reproduction numbers *R*_*0*_ and *R*_*1*_ in the model. This means that in the absence of therapeutic interventions (i.e., anti-COVID-19 drugs and vaccines), successful pandemic control (lowering of R_0_ and R_1_) may be realized by social behavior modifications with enhancement of disease resistance in the susceptible population during quarantine and thereafter. *Hence, release from quarantine should be strongly predicated on continuance and strict adherence to recommended social and health promoting behaviors* (see Table 5a big values).

**Table 5a:** Interpretation of α and ε

| alpha and epsilon |  |
| --- | --- |
| small value | big value |
| highest exposure rate | lowest exposure rate |
| crowding | social distancing |
| no/inadequate PPE | appropriate PPE |
| no handwashing, touching face | proper hand hygiene |
| low disease resistance | high disease resistance |
| natural predisposition | natural resistance |
| malnutrition | good nutrition |
| lack of exercise | physical activity |
| pre existing conditions (e.g. diabetes, hypertension) | good overall health |

The α and ε parameters show significant influence on the number of removed *R* in the population using the Morris Method (global sensitivity analysis). This means that a higher exposure rate (small *α* and *ε*) leads to a faster removal rate. This is akin to accelerating the exposure of the population in order to shorten the duration of the pandemic; however, removal is either by permanent immunity (the object of chickenpox parties) or death. *We strongly caution against hastening the pace of the pandemic because this (1) would overwhelm the health system and (2) may result in unnecessary deaths*.

Additionally, the practical definition of α and ε in terms of their relative values is an important consideration in the model.

In Tables 5a and 5b, the different values of *α* and *ε* are interpreted. Big values for *α* and *ε* (highlighted in green) are ideal control strategies as we want to have the lowest exposure rate, people practice social distancing, etc. Populations who have been exposed, infected, and have recovered may develop immunity and become less vulnerable.

**Table 5b:** Variation of *α* and *ε* values

|  | small epsilon | big epsilon |
| --- | --- | --- |
| small alpha | highest exposure rate | high exposure rate due to behavior of susceptibles |
|  | small spaces between people or no social distancing at all | small spaces between people or no social distancing at all |
|  | low natural immunity | high natural immunity |
|  | high number of vulnerable and/or have pre existing medical condition | low number of vulnerable and/or have pre existing medical condition |
| big alpha | high exposure rate due to behavior of infectives | lowest exposure rate |
|  | have social distancing | have social distancing |
|  | low natural immunity | high natural immunity |
|  | high number of vulnerable and/or have pre existing medical condition | low number of vulnerable and/or have pre existing medical condition |

The reproduction number *R*_*1*_ shows significant sensitivity to α and σ (shown in Tables 1 and 2) suggesting that social distancing, proper hygiene, and contact tracing (for σ) could be effective control strategies for minimizing the spread of the virus post-quarantine. In disease-free equilibrium (DFE), maximizing *α* and *ε* mimics *50%* quarantine, *Q(t) = 0.5*; and as the infectious population increases, the effect of *α* and *ε* decreases.

Against Q-SEIR, Tables 3 and 4 show that with ideal (maximum) values of *α* and *ε* for the post-quarantine scenario (*Q(t) = 1*), the reduction of the infectious population in Quezon City is only by *3.45%*, and in Iloilo Province by *3.88%*. However, as regards peaking dates, there is an earlier surge in the infectious population. This new model projects the peak nine days earlier for Quezon City and 17 days earlier for Iloilo Province.

To improve the model further, age stratification probabilities may be applied as discussed in Bongolan et al. (2020) and Rayo et al. (2020), as well as the saturated treatment rates and Holling Types (II and III) for hospital utilization.

## Data Availability

Data available on request due to privacy/ethical restrictions.

http://ncovtracker.doh.gov.ph/

## APPENDIX

**Fig. 2:**
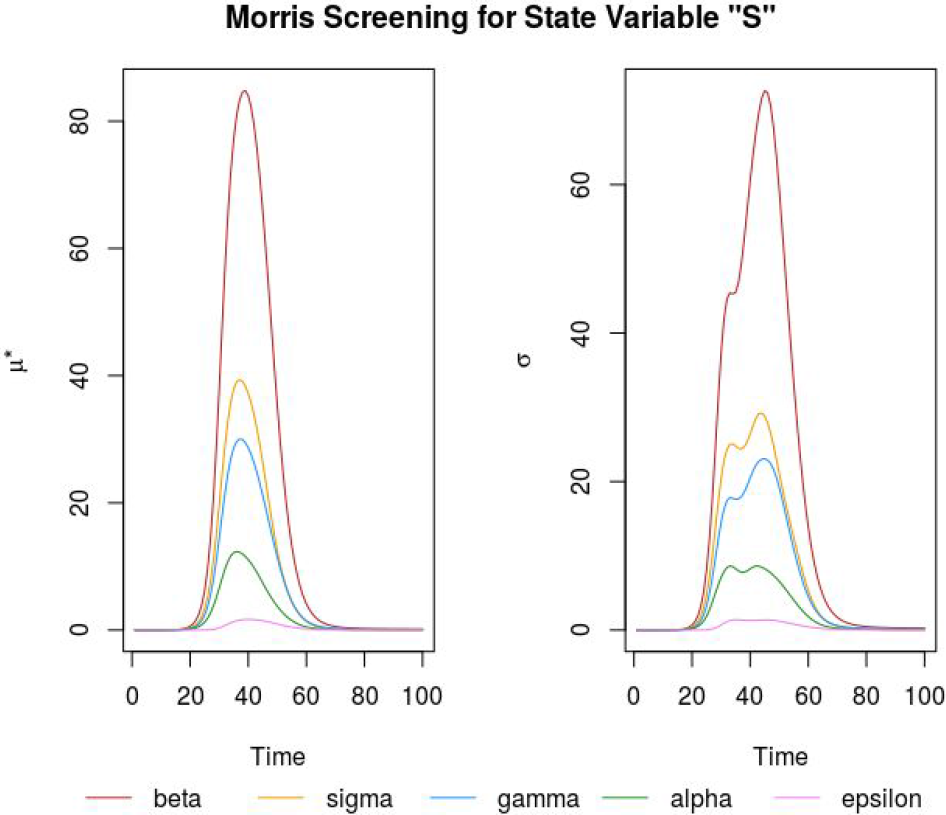
Morris Screening for S

**Fig. 3:**
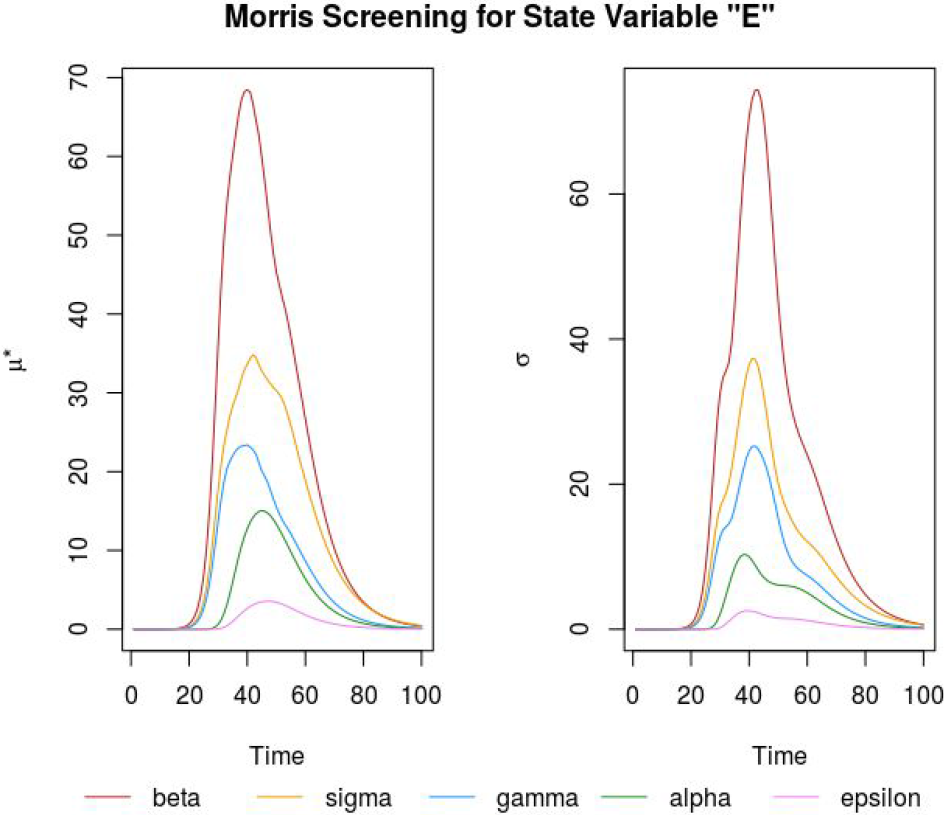
Morris Screening for E

**Fig. 4:**
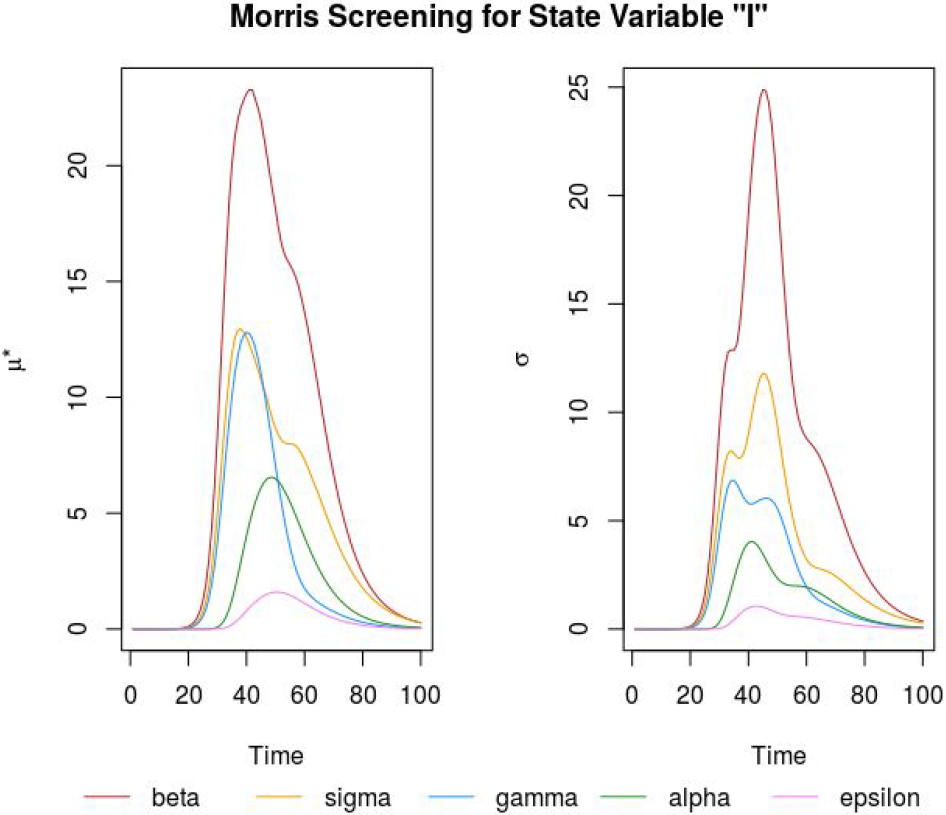
Morris Screening for I

**Fig. 5:**
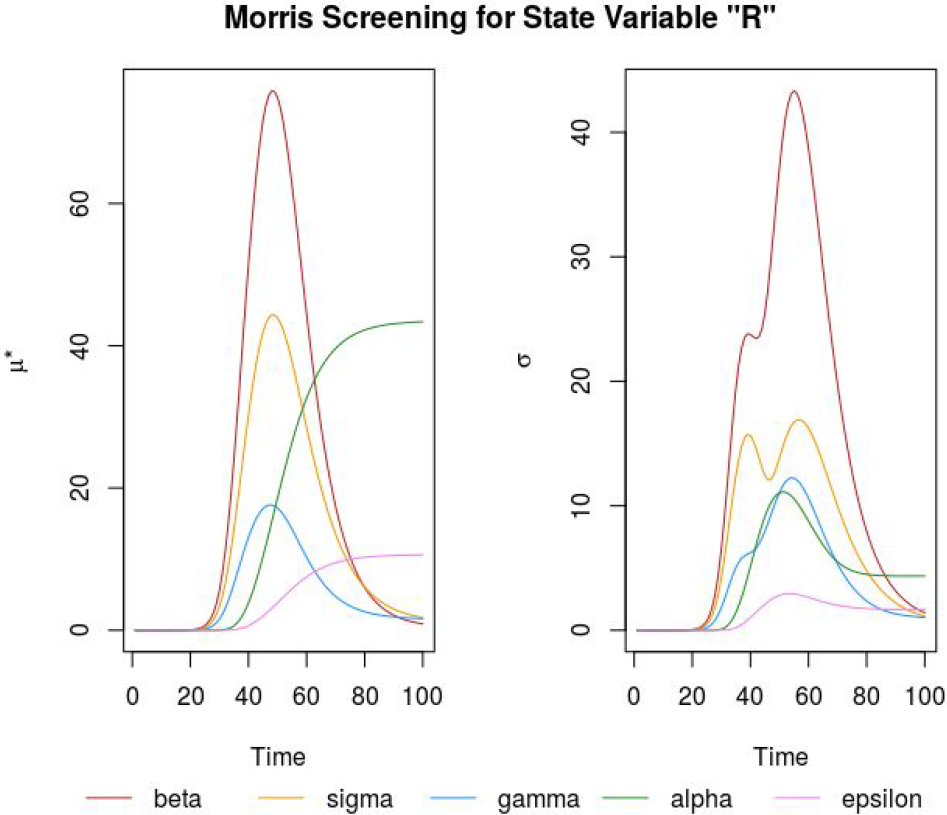
Morris Screening for R

